# Appraising familial prediction of proband outcomes in neurogenetic disorders

**DOI:** 10.64898/2026.05.20.26353681

**Authors:** Shara Reimer, Kathleen Wilson, Lukas S. Schaffer, Isabella Larsen, Melissa Roybal, Srishti Rau, Jyssica Seebeck, Erin Torres, Liv Clasen, Siyuan Liu, Armin Raznahan

## Abstract

**Background:** Gene dosage disorders impact cognition and psychopathology, but outcomes vary widely amongst carriers of the same variant. Recent work has sought to better predict proband outcomes using measures of corresponding traits in family members. However, family-based models have not yet been prospectively quantified across several traits in different genetic disorders, nor evaluated for the precision they afford: both crucial issues for clinical implementation.

**Methods:** In a first test case for these questions, we apply regression analyses to quantify and compare family-based prediction of 12 traits (including IQ, autism- and ADHD-related traits) in 433 individuals from families including a proband with XXY or XYY syndrome (N=93 and 58, respectively).

**Results:** The 12 traits vary substantially in their proband-family associations (0.001<|*r*|<0.55) - with differences emerging between XXY and XYY syndrome. Only two traits also show significant and similar proband-family associations in both aneuploidies, with the greatest concordance found for IQ. A family-based model for IQ prediction in male sex chromosome trisomies significantly reduces error vs. a group mean IQ model (*F* = 7.4, *p* = 0.006), but only in 65% of probands, and with mean error reduction of ∼2 IQ points.

**Conclusions:** Family-based prediction of neuropsychiatric traits in genetic syndromes likely requires trait- and syndrome- specific models. Family models can significantly improve outcome prediction for IQ, but to variable degrees across individuals and with a small mean improvement. By mapping and quantifying these limits, our work helps draft a roadmap for refinement of family-based prediction of proband outcomes in gene dosage disorders.

**Trial Registration:** ClinicalTrials.gov NCT00001246, “89-M-0006: Brain Imaging of Childhood Onset Psychiatric Disorders, Endocrine Disorders and Healthy Controls.” Date of registry: 01 October 1989.

## Background

Advances in psychiatric genetics have identified a growing list of recurrent genetic variants - including chromosomal aneuploidies and sub-chromosomal copy number variations (CNVs) - that substantially increase risk for adverse developmental, cognitive and mental health outcomes [1, 2, 3]. There is consequently great interest in early identification of these risk variants through prenatal [4, 5, 6, 7] and postnatal [8] genetic testing to allow for tailored genetic counseling, early interventions, and patient stratification that could potentially optimize developmental outcomes. However, realizing these gains in precision medicine is complicated by the presence of profound outcome variability across carriers of the same causal variant - a phenomenon well described in numerous intensely studied recurrent variants including CNVs at 22q11 [9] and 16p11 [10], as well as Down syndrome [11] and aneuploidies of the X- and Y-chromosome [12, 13]. The lack of effective tools for accurate prediction of individual-level outcomes in these individually rare- but collectively common- conditions presents a major challenge for tailored genetic counseling and care planning.

One potentially promising approach for better estimating clinical outcomes in carriers of high-impact genetic variants involves harnessing the phenomenon of inter-familial phenotypic correlation [14] to predict variation in neuropsychiatric outcomes across carriers using measures of relevant phenotypes in their unaffected relatives. This correlation captures genetic and environmental sources of trait variance that are shared between the affected individual and their non-carrier family members. Measuring familial features at the time of genetic diagnosis potentially opens a path to prediction of proband outcomes at the earliest pre- and perinatal stages of genetic diagnosis. Despite the theoretical appeal of this idea, it has only been empirically tested in a handful of studies [9, 10, 12, 15, 16] that each focus on only a few outcomes within individual neurogenetic syndromes. Strikingly, there is already emerging evidence across even these sparse studies that the predictability of proband outcomes from family data may vary between different outcomes in a manner that varies yet again between different genetic syndromes. For example, in XYY syndrome, a significant positive relationship between probands and relatives was seen only for two cognition-related outcomes: full-scale intellect quotient (FSIQ) and vocabulary. There were no significant correlations between probands and family members for autism-related or attention-deficit/hyperactivity disorder (ADHD)-related traits [12]. Conversely, in 16p11 deletion, positive proband-family correlations were seen for FSIQ and verbal IQ, as well as autism-related traits (total Social Responsiveness Scale (SRS) score) [10]. Given these inconsistencies, there is a pressing need to formally quantify and compare proband-family predictive relationships in different neurogenetic syndromes for multiple clinical outcomes. The strength and consistency of these predictive relationships across different syndromes and clinical outcomes fundamentally determines whether and how family-based predictions can be used in clinical settings. However, it has been challenging to address this issue due to the rarity of deeply phenotyped families with different neurogenetic disorders. There is a need for quantification of family-based predictions using identical, granular assessment batteries in more than one neurogenetic disorder.

Here, we provide the first prospective appraisal of family prediction models of different traits in different neurogenetic disorders by taking advantage of a rare deep phenotypic family study of different sex chromosome aneuploidies. Specifically, we use a suite of proband-family predictive analyses previously developed and applied to 12 different developmental outcomes in XYY syndrome [12] and extend these to the closely related neurogenetic syndrome – XXY or Klinefelter syndrome (XXY/KS). These two genetic syndromes – characterized by carriage of an extra Y- or X-chromosome in males, respectively - collectively occur in ∼1/400 male births [3, 17, 18] and are both associated with substantially increased risk for diverse neuropsychiatric outcomes including cognitive impairments, autism, ADHD and mood disorders [19]. The direct transfer of identical variables and analytic methods from one sex chromosome trisomy (XYY syndrome) to another (XXY/KS) enables a tight test of whether proband-family predictive relationships are stable across different genetic syndromes and clinical outcomes. To this end, we separately quantify the magnitude of proband-family trait relationships for 12 different traits in XXY/KS, the mean offset in these traits between XXY/KS probands and their family members, and how both properties differ per trait between XXY/KS and XYY. As in our prior study of XYY syndrome, we also test if prediction of proband outcomes in XXY/KS benefits from consideration of socioeconomic and perinatal variables alongside family traits. Our findings specify opportunities for improved prediction of proband outcomes in XYY/KS in particular, and male sex chromosome trisomies in general. Moreover, our explicit comparison of predictive relationships between two different sex chromosome aneuploidies helps to illuminate broader opportunities and challenges in family-based prediction of neuropsychiatric outcomes for carriers of recurrent gene dosage disorders.

## Methods

### Recruitment of probands and family members

Participants this study included 433 individuals from 151 families indexed on a proband with non-mosaic XXY/KS (N=93 families/probands with 167 relatives) or XYY syndrome (N=58 families/probands with 115 relatives) aged between 5 and 25 years. Presence of a sex chromosome aneuploidy was karyotypically confirmed for those 128 probands able to give blood (n=90 XXY/KS, n=38 XYY) or verified by examination of existing clinical genetic reports for the 23 remaining probands (n=3 XXY/KS, n=20 XYY). Participant characteristics [see Additional File 1] and expanded details [see Additional File 2] are provided in additional files. Study participants were recruited through the NIH Clinical Center Office of Patient Recruitment and the Association of X and Y Chromosome Variations. The sole inclusion criterion was presence of a non-mosaic XXY or XYY karyotype in the proband, and - for relatives - being a biological parent of the proband or being a full male (XY) sibling aged 5-26 years. Exclusion criteria for probands included extremely low birth weight, lack of family members participating in the study, and being part of a monozygotic twin pair.

### Measurement of cognition and behavior

Basic demographic, birth history and developmental information were obtained through a clinical interview and sociodemographic questionnaire [MacArthur Scale of Subjective Social Status [20] (Additional File 2)]. General cognitive, verbal and spatial ability were assessed using the FSIQ, vocabulary subtest, and matrix reasoning subtest of an age-appropriate Wechsler scale, encompassing FSIQ. Autism-related traits were measured using the SRS Second Edition questionnaire (SRS-2; [21]), with self-report for adults and parent-report questionnaires for pediatric participants – providing individual-level T-scores for total autism-related traits, social awareness, social cognition, social communication, social motivation, restricted and repetitive behavior (RRB), and social communication and interaction (SCI, a scale which combines the four non-RRB scale scores [social awareness; social cognition; social communication; and social motivation] to yield an overall social score). ADHD traits were measured using the Conners’ Adult ADHD Rating Scales-Self-Report: Long-Version (CAARS-S:L [22]) for adults and the Conners 3-Parent Assessment Report (Conners3 [23]) for pediatric participants – providing individual-level T-scores for inattention and hyperactivity/impulsivity. For all T-scores, higher values reflect greater symptom severity.

### Statistical Analyses

#### Correlation and regression analysis of individual traits between proband and families in XXY/KS

As a basis for comparing proband scores with those for unaffected family members we averaged the scores for all available relatives of each proband into one family-level score. We then used two complementary approaches to quantify the relationship between variation in proband and family scores for each of the 12 measured traits. Pearson correlations estimated the variation in proband scores that could be accounted for by variation in family scores. Ordinary least squares linear regressions were used to model variation in proband scores as a function of family scores as shown below:

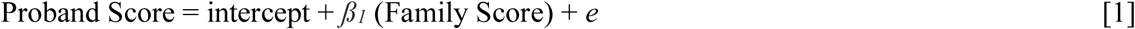

Proband and family scores were standardized across individuals/families using the mean family score, so that (i) the intercept of model [1] estimated the “offset” between proband and family scores at the mean family score, and (ii) the ß*_1_* coefficient for each trait estimated the standard deviation shift in proband score for a 1 standard deviation shift in family score. The intercept therefore captures penetrance of the genetic variant using a family background for reference rather than population norms from unrelated individuals. Testing the intercept value against a null hypothesis of zero provides statistical evidence for an impact of aneuploidy on the trait in question. The slope *ß_1_* coefficients we tested against two alternative null hypotheses: 0 - which tests for evidence of any linear relationship between proband and family scores; and 1 - which tests for linear scaling between family and proband scores (i.e. whether a standard deviation shift in family score is accompanied by a standard deviation shift in proband scores) and therefore also tests whether proband scores show a stable offset across the observed range of family scores. P-values from null hypothesis testing for intercepts and *ß_1_* coefficients were Bonferroni corrected across 12 traits (adjusted *p* significance threshold=0.004).

#### Testing if prediction of proband outcomes is improved by incorporation of socioeconomic and perinatal variables in XXY/KS

Variation in proband outcomes could also relate to variation in environmental factors across families – including socioeconomic (family SES [socioeconomic status] and MacArthur Ladder score) and perinatal factors (proband birth weight and maternal age) [see Additional File 2]. We used the following expanded linear model to test for evidence of this for each proband trait and asked if addition of these variables to family traits can boost prediction of proband outcomes.

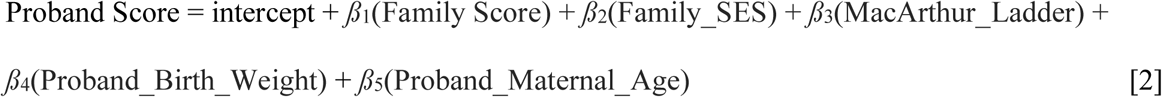

We used an analysis of variance (ANOVA) to compare this full model [2] to the simple univariate model with family score as the sole predictor [1] and thereby test if prediction of proband outcomes benefits from collective consideration of socioeconomic and perinatal factors. P-values from null hypothesis testing for each beta coefficients and for each ANOVA test were Bonferroni corrected across 12 traits (adjusted *p* significance threshold=0.004).

#### Comparing prediction of proband outcomes between XXY/KS and XYY syndrome

Our analytic framework allowed quantitative comparison of proband-family trait relationships between XXY/KS and XYY using the following model for each trait:

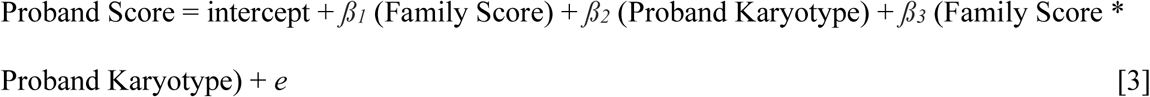

Proband and family scores were standardized using the family mean score for each trait prior to analysis. In these models, Proband karyotype was a binary variable with two levels: XXY/KS and XYY (XYY being the reference level). As such, the intercept estimated the offset between XYY proband scores and family scores at the mean family score; the *ß_1_* coefficient estimated the standard deviation shift in XYY proband score for a 1 standard deviation shift in family score; the *ß_2_* coefficient estimated the difference between proband offsets in XXY/KS vs. XYY at the mean family score; and, the *ß_3_* coefficient estimated the difference in linear relationship between proband-family trait variation in XXY/KS vs. XYY. P-values from null hypothesis testing for each intercept and *ß_1_* coefficient were Bonferroni corrected across 12 traits (adjusted *p* significance threshold=0.004).

#### Quantifying the reduction of error afforded by use of family data in prediction of proband outcomes

The analyses above highlighted IQ as the proband trait that was most robustly predicted by family scores for both XXY/KS and XYY. We compared the accuracy of proband IQ prediction in male sex chromosome trisomies using (i) predicted IQ based on karyotype-specific regression model with family IQ (error estimated as the signed residual between observed IQ and predicted IQ from single model [3] – henceforth “family model error”) vs. (ii) predicted IQ from the group mean (error estimated as the signed difference between observed proband IQ and the mean IQ of their respective karyotype group (henceforth “group mean model error”). To formally test whether the family model provided more precise predictions, we used a Levene’s test to compare the variance of the prediction errors from the family and group mean models, as a smaller variance indicates greater predictive accuracy. We also calculated the absolute difference between family model and group mean error values for each proband and then averaged these by karyotype to estimate the mean reduction in FSIQ prediction error provided by use of the family model.

All data analyses and visualizations were performed using the following packages: *purrr* [24]*, dplyr* [25]*, magrittr* [26]*, ggplot2* [27]*, viridis* [28], *tidyverse* [29], *MASS* [30]*, sandwich* [31], *lmtest* [32], *plyr* [33], and *car* [34] in RStudio using R version 4.5.0 [35]

## Results

### Participant Characteristics

Participant characteristics are summarized in Additional File 1 and detailed in in Additional File 2. After Bonferroni correction, Welch two-sample t-tests revealed a significant difference in mean family value by karyotype for only one trait: SRS social awareness (*p* < 0.0001). There were significant differences that did not survive Bonferroni correction for vocabulary (*p* = 0.02), SRS total (*p* = 0.04), SRS communication (*p* = 0.01), and SCI (*p* = 0.02). The mean scores were within the normal range for both family cohorts, and the difference between cohorts was minimal (XYY family score mean – XXY family score mean, in points: social awareness = 5.4, vocabulary = - 0.7, SRS total = 2.0, communication = 2.6, SCI = 2.2).

### Proband-Family Trait Relationships in XXY/KS

Results from correlation and regression analysis of proband-family trait relationships in XXY/KS are detailed in ***Table 1***. Proband-family trait correlations were generally positive (mean correlation across traits: 0.32, range: 0.01 to 0.54). The three strongest positive correlations were seen for FSIQ and vocabulary from the Wechsler scales (*r* = 0.54 and 0.45, respectively) and the SRS SCI subscale (*r* = 0.43). Correlations were near zero for inattention and hyperactivity/impulsivity.

**Table 1.** Results of analyses interrelating variation in proband and family scores for 12 traits in XXY/KS.

| Variable | Correlation Coefficient (*r*) | Adjusted R<sup>2</sup> | Intercept | Standardized Intercept | *Intercept p* | Slope | Slope *p* (vs. 1) | Slope *p* (vs. 0) |
| --- | --- | --- | --- | --- | --- | --- | --- | --- |
| FSIQ | 0.54 | 0.28 | -15.51 | -1.03 | < 0.0001* | 0.57 | < 0.0001* | < 0.0001* |
| Vocab | 0.45 | 0.2 | -3.17 | -1.06 | < 0.0001* | 0.61 | 0.002* | < 0.0001* |
| MR | 0.32 | 0.09 | -1.73 | -0.58 | < 0.0001* | 0.32 | < 0.0001* | 0.002* |
| SRS Total | 0.41 | 0.16 | 11.43 | 1.14 | < 0.0001* | 0.74 | 0.13 | < 0.0001* |
| Soc Awr | 0.23 | 0.04 | 10.3 | 1.03 | < 0.0001* | 0.39 | 0.0006* | 0.025 |
| Soc Cog | 0.36 | 0.12 | 11.9 | 1.19 | < 0.0001* | 0.71 | 0.14 | 0.0004* |
| Soc Comm | 0.30 | 0.08 | 12.1 | 1.21 | < 0.0001* | 0.55 | 0.017 | 0.004* |
| Soc Mot | 0.41 | 0.16 | 6.94 | 0.69 | < 0.0001* | 0.56 | 0.001* | < 0.0001* |
| RRB | 0.29 | 0.07 | 8.37 | 0.84 | < 0.0001* | 0.58 | 0.04 | 0.005 |
| SCI | 0.43 | 0.17 | 11.9 | 1.19 | < 0.0001* | 0.75 | 0.14 | < 0.0001* |
| Inatt | 0.12 | 0.004 | 19.35 | 1.94 | < 0.0001* | 0.15 | < 0.0001* | 0.25 |
| Hyp Imp | 0.01 | -0.01 | 11.46 | 1.15 | < 0.0001* | 0.01 | < 0.0001* | 0.95 |

*Note.* FSIQ = Full-Scale IQ; Vocab = Vocabulary; MR = Matrix Reasoning; SRS Total = Social Responsiveness Scale (SRS) Total T-Score; Soc Awr = SRS Social Awareness T-Score; Soc Cog = SRS Social Cognition T-Score; Soc Comm = SRS Social Communication T-Score; Soc Mot = SRS Social Motivation T-Score; RRB = SRS Restricted Interests and Repetitive Behavior T-Score; SCI = SRS Social Communication and Interaction T-Score; Inatt = Conners Inattention T-Score; Hyp Imp = Conners Hyperactivity/Impulsivity T-Score. Bold *p* values are significant, and bold *p* values with an asterisk are significant and survive Bonferroni correction.

Linear regression analyses (***Table 1***) confirmed the presence of a statistically significant non-zero linear relationship between proband and family scores for almost all of the 12 traits considered, except inattention, hyperactivity/impulsivity, social awareness, and restricted/repetitive behaviors. For all traits with significantly non-zero *ß_1_* coefficients, *ß_1_* coefficients were below 1 (mean: 0.60, range: 0.32-0.75) – indicating a consistent tendency to sub-linear scaling between proband and family scores, in which a standard deviation increase in family score was accompanied by a smaller estimated increase in proband score. However, of the traits with a *ß_1_* coefficient significantly different from 0, *ß_1_* coefficients were only statistically distinguishable from 1 for FSIQ, Vocabulary, Matrix Reasoning, and SRS Social Motivation scores - providing statistical evidence that the offset of these scores in probands relative to their families varies across the range of family scores. In contrast, *ß_1_* coefficients for SRS Total, Social Cognition, Social Communication, and SCI scores were not statistically distinguishable from 1 - implying a stable offset of proband scores relative to family background across the range of observed family scores. For all traits, intercept values estimated the average offset in probands relative to family members at the mean family score. Converting these intercept values to standardized units through division by the scale’s respective standard deviation (***Table 1***) revealed a mean absolute offset of 1.09 across traits – with prominent offsets for overall autism-related traits (SRS Total Score standardized offset = 1.14), several subscales of autism-related symptomatology (especially social communication, standardized offset = 1.21) and ADHD-related symptomatology (all standardized offsets >1), and IQ (standardized offset = -1.03, driven more by impairments of vocabulary than matrix reasoning). ***Figure 2*** illustrates proband-family score interrelationships for selected traits (plots for all traits shown in ***Fig S1***): IQ and vocabulary (significant proband-family relationships with proband offsets that vary by family score); total autism-related symptomatology (a significant proband-family relationship without statistical evidence for a varying offset by family score); and, hyperactivity/impulsivity (no apparent proband-family relationship with a significant proband offset at mean family score).

**Figure 1:**
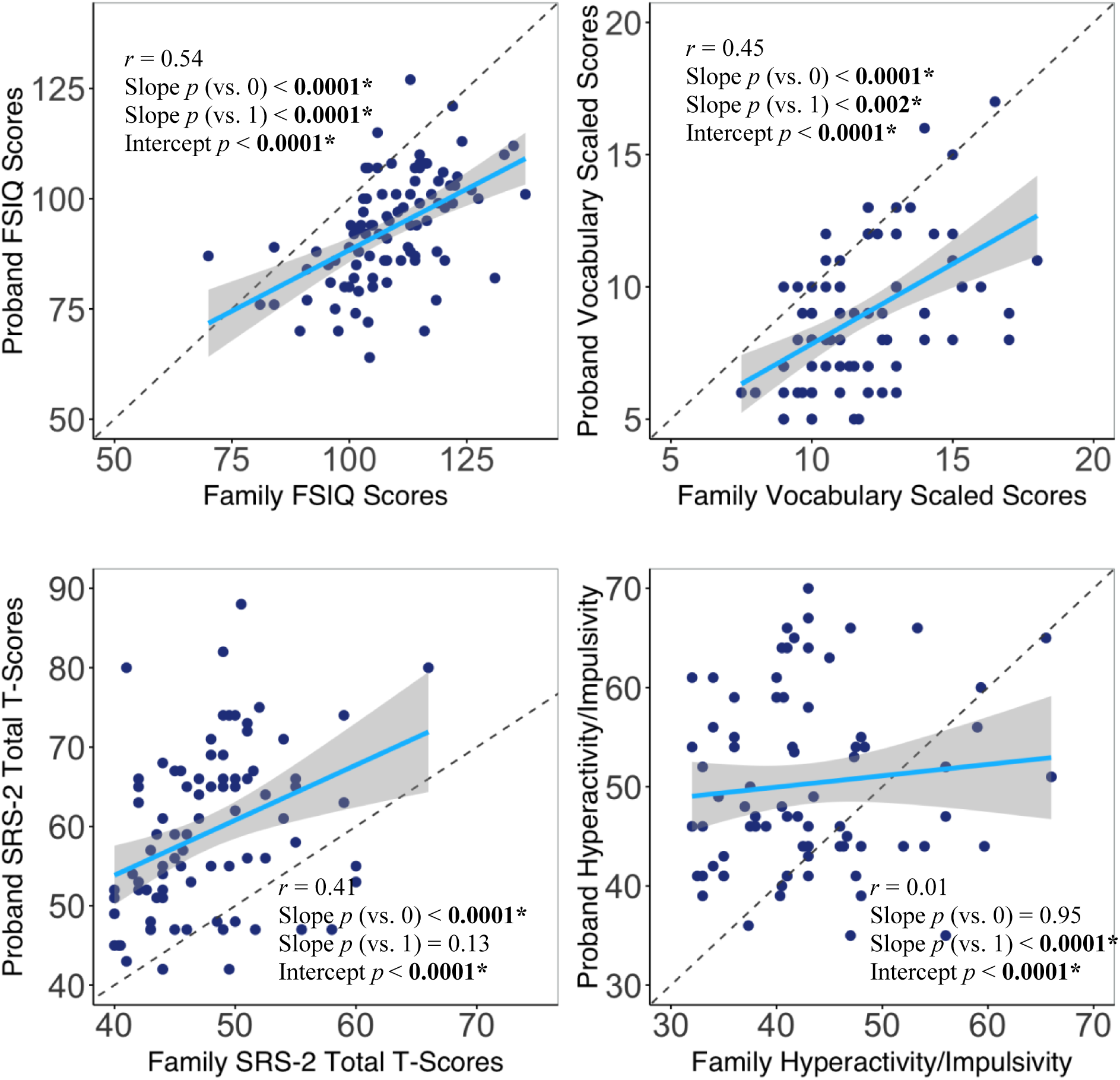
Scatterplots for selected proband-family trait relationships in XXY/KS. In each scatterplot, points are individual proband-family units, dotted gray lines show the x=y identity line and solid blue lines show the least squares regression fit line predicting proband scores (y-axis) from family scores (x-axis).

**Figure 2:**
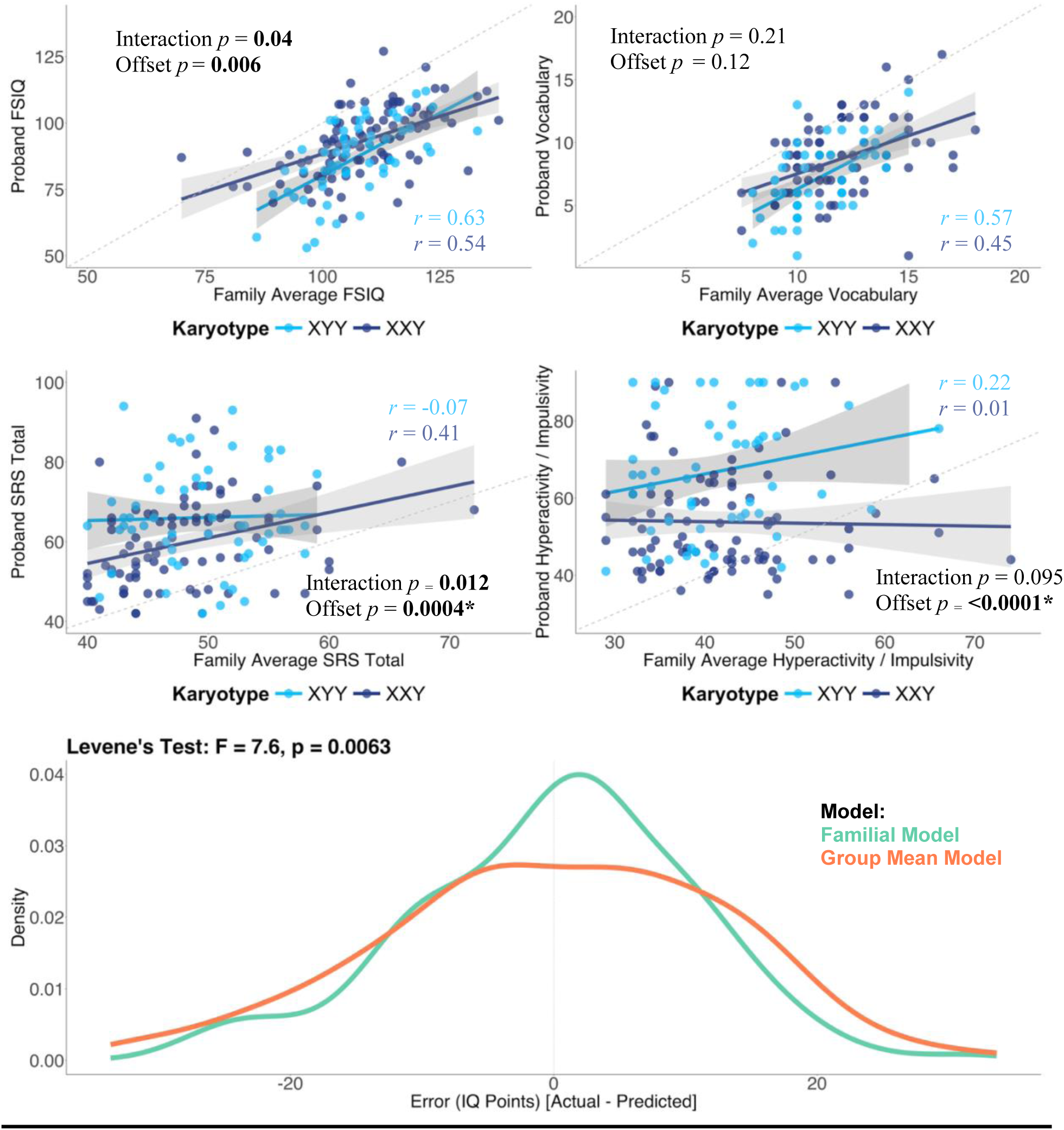
Comparing family modeling of proband outcomes between XXY/KS and XYY analyses. *Top*: In each scatterplot, points are individual proband-family units. Solid bluelines show the least squares regression fit line predicting proband scores (y-axis) by family scores (x-axis) for XXY/KS (dark blue) and XYY syndrome (light blue). *Bottom*: Density plot showing the distribution of errors in proband FSIQ prediction from a family model (green) as compared to a karyotype group mean model (orange). The variance in error scores is significantly less for the family vs. mean model (*F* = 7.4, *p* = 0.006), indicating improved prediction of outcomes using family data.

Of note, there were no traits that could be better predicted in the proband by additional consideration of socioeconomic and perinatal variables (all ANOVA F-tests p>0.05 for comparison model [2] and [1] above).

### Comparing family-based prediction of proband outcomes in XXY/KS vs. XYY Syndrome

We next sought to compare family-based prediction of proband scores in XXY/KS to those in XYY by combining both cohorts and including karyotype as a main and interactive effect when modeling variation on proband scores by variation in family scores (i.e. model [3] above). These analyses [see Additional file 3] found a general pattern of more severe impairments relative to family background for XYY than XXY/KS, and revealed that the regression slopes predicting proband from family scores were statistically significantly different between XXY/KS and XYY after Bonferroni correction for social awareness and social motivation. However, for 4 of the remaining traits – SRS total, social communication, inattention and hyperactivity/impulsivity – variation in proband scores could not be significantly predicted by variation in family scores. Thus, of the 12 traits examined, only 6 showed proband-family score relationships that were both (1) not statistically distinguishable between XXY/KS and XYY syndrome, and (2) statistically significant across all male sex chromosome trisomy families combined: FSIQ, vocabulary, matrix reasoning, social cognition, RRB, and SCI. Of these 6 traits, family-proband relationships were notably strongest for FSIQ and vocabulary. Examples of the diverse proband-family models in XXY/KS and XYY are shown as scatterplots in **Fig 2**.

### A Combined Model for Family-based Prediction of Proband IQ in Male Sex Chromosome Trisomies

Of all 12 measured traits, the analyses above highlighted FSIQ as providing a promisingly strong predictive relationship between family and proband scores across both XXY/KS and XYY. We therefore focused on this trait as a test-case for developing a combined predictive model for both male sex chromosome trisomies and quantifying how well this model reduced error in prediction of individual-level FSIQ relative to the alternative of using mean IQ of the proband group. To achieve this, we modeled proband FSIQ as a function of mean family FSIQ using model [3] across all 151 sex chromosome aneuploidy families (***Table 3*, *Figure 2***). We recorded the difference between observed proband FSIQ and FSIQ as predicted from this family model (henceforth “family model error”). As a comparison, we also computed the difference between observed proband FSIQ and the mean FSIQ of their respective karyotype group (“group mean model error”). We then compared family model and mean model error scores, pooled across karyotype groups.

Family model error scores were more tightly centered around zero than group mean model error - scores indicating a general tendency towards improved prediction of proband FSIQ (Levene’s test: *F* = 7.6, *p* = 0.006; **Figure 2**). However, the mean difference in error between family- and group mean-models across all probands using model [3] was 2.44 (XXY/KS: 2.35; XYY: 2.57) and the range in error differences across individuals was large (-15.1 to 23.6; XXY/KS: -15.1 to 17.4; XYY: -13.0 to 23.6). The family model provided a better prediction of IQ than the group mean model in 65% of probands (XXY/KS: 69%; XYY: 54%).

## Discussion

This study provides a tight comparison to test both the strength and universality of proband-family relationships for diverse neuropsychiatric traits across two genetic disorders. Our findings have important implications for the potential usage of family-based models to improve prediction of person-level clinical outcomes in gene dosage disorders.

As a basis for comparison of family prediction in XXY/KS and XYY, we present the first analyses of family-based predictions in XXY/KS. These analyses yield several insights of note. First, they provide estimates for the penetrance of XXY/KS using family scores rather than population normed scores as a reference – helping to control against any sources of ascertainment bias involving genetic and environmental factors that vary across families. Thus, the observed shifts in mean proband score relative to expectations from the mean family background - quantified by the intercepts from proband-family regression models - better approximate the true population-based penetrance of XXY/KS than would otherwise be possible using data from clinical proband cohorts. Even with this improved control for ascertainment bias, we still observe large mean impacts of XXY/KS with standardized effect sizes exceeding 1.5 on several neuropsychiatric traits. Second, the regression slopes modelling proband outcomes from family scores are significantly below 1 for most traits – meaning that the estimated deviation of mean proband scores from family background varies as a function of family scores. This observation is as expected from classical quantitative genetics [36, 37] and has important implications for any envisaged implementation of family-based prediction models in genetic counselling and psychoeducation (given that proband offsets will be greater in the context of higher family scores). Third, as seen before in XYY syndrome [12], we find that the strength of proband-family correlations also varies greatly across traits in XXY/KS, where it ranges from *r* of 0.01 to 0.54. This variability rules out the utility of family-based prediction of proband outcomes for several traits in XXY/KS (especially those related to ADHD), but it opens the door to potential utility for others (especially general cognitive ability). As in our past study of XYY syndrome [12], FSIQ showed the most significant proband-family correlation, with a similar correlation slope to those seen in several other gene dosage disorders [9, 10, 15, 16]. This observed slope value aligns perfectly with theoretical expectations [36, 37] that the regression slope for prediction of an offspring’s traits from the bi-parental mean should approximate the trait’s heritability (which is ∼0.6 for FSIQ in the general population [38]). Moreover, similar to findings in studies of XYY syndrome [12], XXY/KS siblings [16], and 16p11.2 deletion disorder [10] and in keeping with published heritability estimates [39], there was a greater correlation between probands and family members for verbal versus non-verbal subcomponents of FSIQ.

In addition to exploring the potential for family-based prediction models in XXY/KS, a second major goal of our study was to use the comparison of XXY/KS and XYY syndrome as a formal test for the stability of family-based prediction models across different genetic cohorts. This test has important implications for the implementation of family-based prediction models in clinical settings. If different traits need different proband-family regression models in different genetic syndromes, then this poses a much more complex and harder-to-scale implementation scenario that one where one model can be applied in several trait/syndrome contexts. We find that of the 12 traits studied, only 3 show a significant proband-family association in both XXY/KS (current study) and XYY [12] (FSIQ, vocabulary and social awareness), and one of these traits (social awareness) has a proband-family model regression slope that is significantly different between XXY/KS and XYY. This striking variability in proband-family associations between two closely related gene dosage disorders studied with a harmonized clinical research protocol indicates that the prior observation of variable prediction models across separate studies of previous traits and genetic syndromes [9, 10, 12, 15, 16] was unlikely to reflect purely methodological heterogeneities. Rather, our current findings strongly suggest that any envisaged clinical implementation of proband-family prediction models will indeed demand bespoke models for each trait-syndrome context – with profound implementations for scalability.

Although most of the proband-family relationships we examine herein appear to be problematically variable across different traits and syndromic contexts, we do find relatively strong and stable proband-family associations for FSIQ in XXY/KS and XYY which are of a similar magnitude to those seen in other gene dosage disorders and the general population [9, 10, 12, 15, 16, 40]. Although FSIQ is just one developmental outcome of many, and its measurement and meaning are not without controversies [41], it is impacted in most neurogenetic syndromes and does show good predictive validity for diverse aspects of health, wellbeing, and adaptive function [42]. As such, improved prediction of proband FSIQ from family measures could have clinical utility in genetic counseling and care planning. We find that a combined family-based prediction model for both male sex chromosome trisomies can significantly reduce error in prediction of proband outcomes relative to the default prediction from group mean IQ. However, the mean magnitude of error reduction is small with a wide range, such that the family model provides a worse estimate than the observed group mean in ∼1/3 of probands.

Our findings should be considered in light of several caveats and limitations. First, as for our past study of XYY syndrome, the XXY/KS families included in this study were not recruited through population-based sampling and are therefore liable to potential ascertainment bias effects. We note however that estimating the penetrance of XXY/KS and XYY effects within a family-based study design helps to mitigate some sources of ascertainment bias relative to the majority of natural history studies in clinical cohorts which lack accompanying trait measures from family members. Second, our study is cross-sectional in design and cannot account for age-varying proband-family interrelationships. Third, some of our measures include rating scales with different raters, as parents answered for probands and siblings, but parent data was self-reported. There could be rater effects which our study did not directly model but which could deflate estimates of proband-family trait correlations. Fourth, our study design is purely observational and cannot disambiguate the many potential drivers for proband-family trait correlations such as shared genetic and environmental factors or bidirectional causal relationships between outcomes in probands and their family members. Finally, while our sample size is large relative to prior family-based studies of gene dosage disorders [10, 12, 15]. and we provide the first direct comparison of proband-family correlations for multiple traits between two different genetic disorders, future work will need to expand the number of families, traits and genetic disorders studied.

### Conclusions

Notwithstanding the above limitations, our study harnesses deep phenotypic data in families with two different sex chromosome aneuploidies to clarify important translational questions around potential implementation of family-based predictive models in clinical practice. Our findings suggest that many clinical outcomes will need unique predictive models tailored for different trait- syndrome contexts. Although FSIQ shows relatively large and stable proband-family relationships between XXY/KS, XYY and other gene dosage disorders, we show that the individual-level accuracy this affords for FSIQ prediction is highly variable and - on average - weak. These findings help to motivate and focus future work on family-based prediction of proband outcomes in neurogenetic disorders. Future work should explore the potential for boosting the predictive power of family models through (i) multivariate models that simultaneously encompass multiple cognitive and behavioral phenotypes, and (ii) incorporation of additional genotypic, environmental and biological features. Addressing these questions will be essential for fully appraising the potential efficacy of family-based prediction models in neurogenetic disorders.

## List of Abbreviations

CNV: copy number variation
FSIQ: full-scale intellect quotient
ADHD: attention-deficit/hyperactivity disorder
SRS: Social Responsiveness Scale
RRB: restricted and repetitive behavior
SCI: social communication and interaction
CAARS-S:L: Conners’ Adult ADHD Rating Scales- Self-Report: Long-Version
SES: socioeconomic status
ANOVA: analysis of variance

## Declarations

### Ethics approval and consent to participate

All participants gave consent or assent for study procedures, and all procedures were approved by the National Institutes of Health Institutional Review Board.

### Consent for publication

Not applicable.

### Availability of data and materials

The data are registered with ClinicalTrials.gov under the name “89-M-0006: Brain Imaging of Childhood Onset Psychiatric Disorders, Endocrine Disorders and Healthy Controls.”

### Competing interests

The authors declare that they have no competing interests.

### Funding

This study was fully supported by the intramural research program of the National Institute of Mental Health (NIMH) (NIH Annual Report Number: ZIAMH002949; Protocol: 89-M-0006; ClinicalTrials.gov Number: NCT00001246). The content is solely the responsibility of the authors and does not necessarily represent the official views of the National Institutes of Health.

### Authors’ contributions

SRe conceptualized the project, cleaned and analyzed data, and drafted the manuscript. KW conceptualized much of the analytical framework used in the project. LS, IL, MR, SRa, JS, and ET were major contributors to data collection for the project. LC was a major contributor to preparing and cleaning data for the project. SL served as the statistical expert for the project. AR was a major contributor in conceptualizing the project and writing the manuscript. All authors reviewed the manuscript.

## Acknowledgements

The authors thank all participants and families that participated in this study.

## Authors’ information

The contributions of the NIH authors (SRe, MR, SRa, JS, ET, LC, SL, and AR) are considered Works of the United States Government. The findings and conclusions presented in this paper are those of the authors and do not necessarily reflect the views of the NIH or the U.S. Department of Health and Human Services.

## Additional Files

Name: Additional File 1 Format: .xlxs

Title: Participant Characteristics.

Description: This table displays summary statistics for each measured trait for each category of participant. FSIQ is reported as a standard score. Vocabulary and matrix reasoning are reported as scaled scores. All other measures are reported as T-scores. N shows the number of participant values (XXY proband/mother/father/sibling, XYY proband/mother/father/sibling) for each measure. The SRS-2 subscales have the same N as the SRS-2 Total Score. The ADHD measures have the same N for both ADHD-traits. FSIQ=Full-Scale Intelligence Quotient, SRS-2=Social Responsiveness Scale Second Edition. SRS-2 Awareness=SRS-2 Social Awareness, SRS-2 Cognition=SRS-2 Social Cognition, SRS-2 Communication=SRS-2 Social Communication, SRS-2 Motivation=SRS-2 Social Motivation, SRS-2 RIRB=SRS-2 Restricted Interests and Repetitive Behaviors, SRS-2 SCI=SRS-2 DSM-5 Social Communication and Interaction.

Name: Additional File 2 Format: .xlxs

Title: Extended demographic information for study participants.

Description: This table shows sociodemographic information for study participants. For Family SES, a lower score indicates a higher SES. All demographic data was self-reported by parents.

Name: Additional File 3 Format: .xlxs

Title: Comparison of proband-family prediction models between XXY and XYY syndromes. Description: This table shows a comparison between the proband-family relationships seen for a variety of traits in XXY and XYY syndromes. FSIQ = Full-Scale IQ; Vocab = Vocabulary; MR = Matrix Reasoning; SRS Total = Social Responsiveness Scale (SRS) Total T-Score; Soc Awr = SRS Social Awareness T-Score; Soc Cog = SRS Social Cognition T-Score; Soc Comm = SRS Social Communication T-Score; Soc Mot = SRS Social Motivation T-Score; RRB = SRS Restricted Interests and Repetitive Behavior T-Score; SCI = SRS Social Communication and Interaction T-Score; Inatt = Conners Inattention T-Score; Hyp Imp = Conners Hyperactivity/Impulsivity T-Score. Bold *p* values are nominally significant, and bold p values with an asterisk survive Bonferroni correction.

---

**Supplementary Figure 1:**
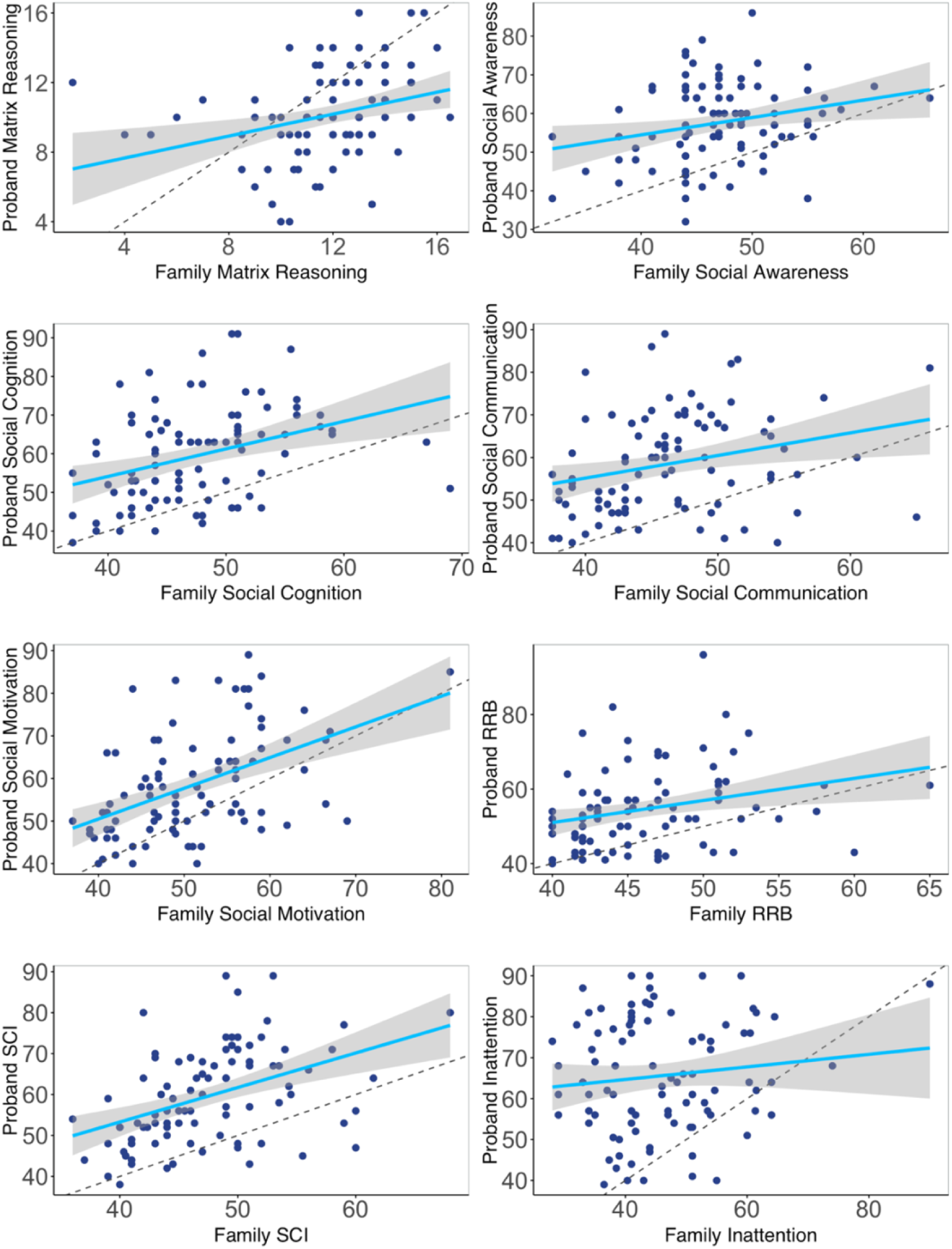
Scatterplots for additional proband-family trait relationships in XXY/KS. In each scatterplot, points are individual proband-family units, dotted gray lines show the x=y identity line and solid blue lines show the least squares regression fit line predicting proband scores (y-axis) from family scores (x-axis).

**Supplementary Figure 2:**
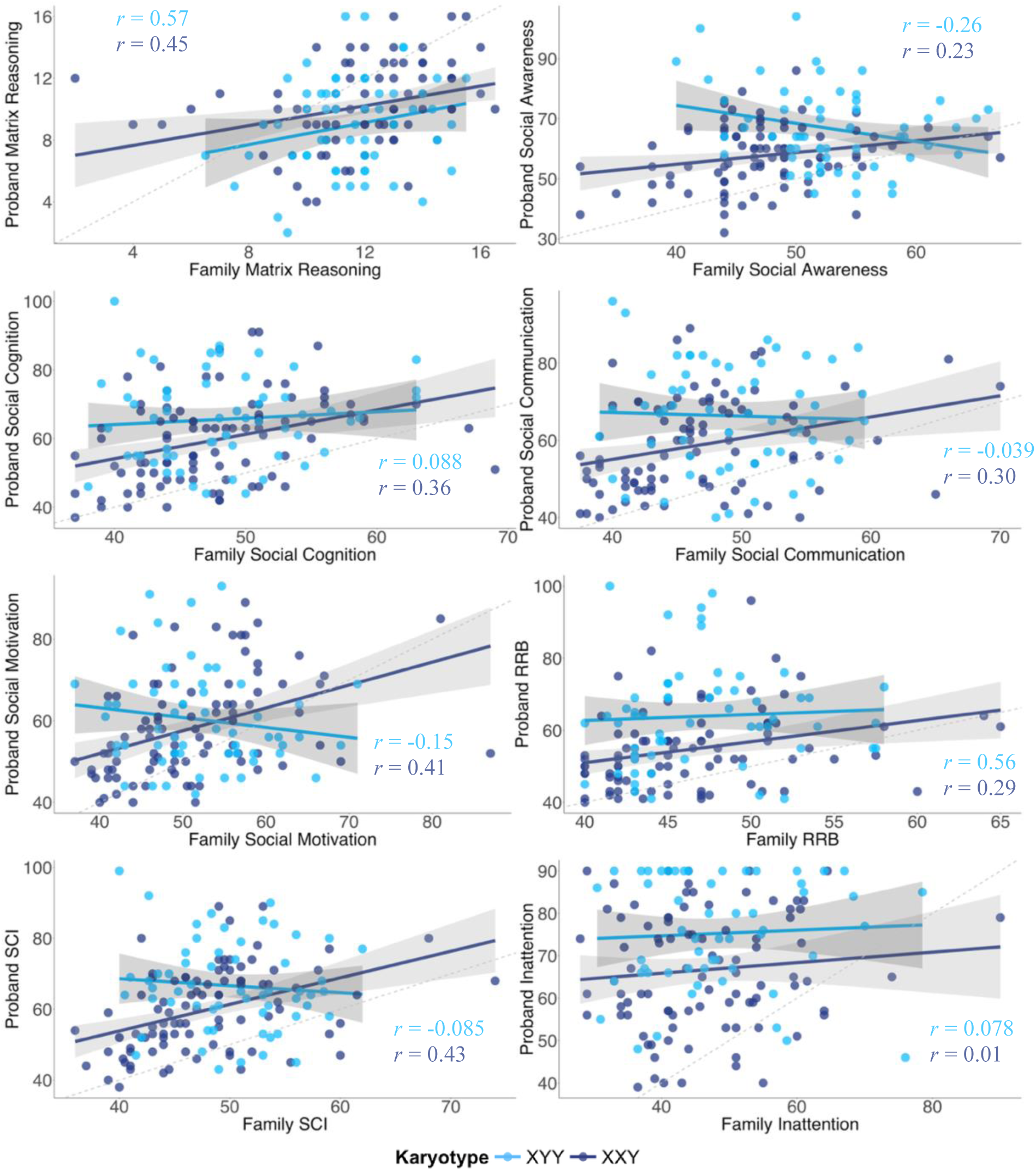
**Additional combined XXY/XYY analyses**. In each scatterplot, points are individual proband-family units. Solid bluelines show the least squares regression fit line predicting proband scores (y-axis) by family scores (x-axis) for XXY/KS (dark blue) and XYY syndrome (light blue).

## Notes

### Competing Interest Statement

The authors have declared no competing interest.

### Clinical Protocols

https://clinicaltrials.gov/study/NCT00001246

### Author Declarations

The IRB of the National Institutes of Health gave ethical approval for this work.

